# GARD is a pan-cancer predictor of radiation therapy benefit

**DOI:** 10.1101/2020.12.19.20248484

**Authors:** Jacob G. Scott, Geoffrey Sedor, Patrick Ellsworth, Jessica A. Scarborough, Kamran Ahmed, Daniel E. Oliver, Steven A. Eschrich, Javier F. Torres-Roca, Michael W. Kattan

## Abstract

**Background:** Despite advances in cancer genomics, radiation therapy (RT) is still prescribed based on an empiric one-size-fits-all paradigm. Previously, we proposed a novel algorithm using the genomic adjusted radiation dose (GARD) to personalize RT prescription dose based on the *biological effect* of a given physical RT dose, calculated using individual tumor genomics. We hypothesize that GARD will reveal interpatient heterogeneity associated with opportunities to improve outcomes compared to physical RT dose alone. To test this hypothesis, and the GARD-based RT dosing paradigm, we performed a pooled pan-cancer analysis in 11 separate clinical cohorts of 1,615 unique patients with 7 different cancer types that represent all available cohorts with the data required to calculate GARD, together with clinical outcome.

**Methods:** Using 11 previously-published datasets of cancers including breast, head and neck, non-small cell lung, pancreas, endometrium, melanoma and glioma, we defined two clinical endpoints: (i) time to first recurrence and (ii) overall survival, comprising 1,298 (982 +RT, 316 -RT) and 677 patients (424 +RT, 253 -RT), respectively. We used Cox regression stratified by cohort to test association between GARD and outcome with separate models using RT dose and sham-GARD for comparison. Interaction tests between GARD and treatment (+/- RT) were performed using the Wald statistic.

**Results:** Pooled analysis of all available data reveal that GARD as a continuous variable is associated with recurrence (HR = 0.982, CI [0.970, 0.994], p = 0.002) and survival (HR = 0.970, CI [0.953, 0.988], p = 0.001). The interaction test revealed the effect of GARD on survival depends on whether or not that patient received RT (Wald statistic: p=0.011). Physical RT dose and sham-GARD were not significantly associated with either outcome.

**Conclusions:** The biologic effect of radiation therapy, as quantified by GARD, is significantly associated with recurrence and survival for those patients treated with radiation: it is predictive of RT benefit; and physical RT dose is not. We propose integration of genomics into radiation dosing decisions, using a GARD-based framework, as the new paradigm for personalizing RT prescription dose.

## Introduction

Radiation therapy (RT) is the oldest and single most utilized cytotoxic therapy in oncology and is responsible for 40% of all cancer cures.^1,2^ There have been innumerable technological advances over RT’s *>* 100 year history, allowing for better *anatomic RT dose targeting*. There have not, however, been such advances in the *physical RT dosing paradigm*, where we still prescribe one-size-fits-all uniform doses to all patients with a given cancer diagnosis using a protracted-fractional method, based on experiments first done on rams and rabbits more than 80 years ago.^3^ Physical RT dose is prescribed based on energy absorbed by tissue (measured in Gray) and historically the field has recognized that the physical interaction of radiation with tissue results in a *biological effect* (e.g. DNA damage, clinical tumor response, carcinogenesis).^4^ However, there is significant interpatient heterogeneity in the biological effect of a given physical RT dose – *patients we treat uniformly do not have a uniform response*. We have shown previously that this difference in radiation induced biological effect can be quantified at the patient level using tumor genomics, and subsequently, that it can be modulated by the treating radiation oncologist.^5,6^ While the sequencing of the human genome changed the therapeutic paradigm in medical oncology, the same is not yet true for radiation oncology, where an empiric paradigm is still utilized to prescribe one-size-fits-all RT dosing: that is, all patients with a given cancer type receive the same prescription RT physical dose.

The clinical heterogeneity of radiation response, even within disease groups, is well established. Further, that this heterogeneity is at least partly driven and influenced by changes in the tumor genome is now accepted – indeed, large-scale classification efforts have been performed to understand these differences.^7–9^ There have also been several efforts to understand surrogate genomic or clinical metrics for individual patient’s resistance to radiation^10^, including, but not limited to HPV status in head and neck cancer^11^, as well as imaging-based studies in both CT-based radiomics^12^ and functional imaging.^13^ Further, several groups have worked together to create signatures indicative of patient groups who can have radiation omitted.^14^ In addition there have been efforts in developing early markers of persistent microscopic disease as circulating tumor DNA (ctDNA).^15^ None of these efforts, however, considers the explicit relationship between intrinsic tumor genomics and physical RT dose – the standard, and fundamental, parameter in radiation therapy.

We previously introduced the gene expression-based radiosensitivity index (RSI), a biomarker of tumor radiosensitivity, which we, and others, have validated in multiple cohorts spanning various disease sites by dichotomizing patients as either radiosensitive or radioresistant.^5,6,16–28^ Subsequently, we developed the Genomic Adjusted Radiation Dose (GARD), which integrates RSI and physical RT dose to *quantify the biological effect of a given physical RT dose* in an individual patient.^4,6^ This created the first opportunity to understand the relationship between intrinsic tumor radiation sensitivity and the biological effect of a given RT dose. Further, GARD allows a clinician to dose a patient’s tumor to a desired biological effect (measured in GARD units), permitting personalization of physical RT dose: a new RT prescription paradigm in which information about physical RT dosing is enriched by a genomic dimension.

To validate the GARD-based RT dosing paradigm, we performed a pooled, pan-cancer analysis to compare disparate outcomes and disease sites in which we utilize all available clinical datasets with enough information to calculate GARD.^29^ In this manuscript, we analyze the association of biologic effect, quantified by GARD, with recurrence and survival across 7 disease sites, encompassed by 11 separate cohorts, totaling 1,615 patients.

## Material and Methods

### Clinical Cohorts and Endpoints

We collected a series of 11 previously-published datasets for which we had all information to calculate GARD (gene expression via microarray, physical RT dose and dose per fraction) and clinical outcome. GARD was calculated as previously reported, scaling RSI within each cohort (for details on this calculation and the genes involved in RSI, see **Supplemental Equations 1 and 2**).^6^ These cohorts, summarized in **Table 1**, include patients treated with RT for breast cancer (BC), including triple negative breast cancer (TNBC),^21,22,30^ glioma,^17^ pancreatic,^23^ endometrial,^27^ melanoma^24^, head and neck^16^ (HNC) and non-small cell lung^6^ cancers (NSCLC). In addition, it includes patients that were not treated with RT as a negative control, to determine GARD is specific for RT-treated patients. While these cohorts are imperfect, they represent all available cohorts with the information available to calculate the variable of interest: and taken together represent a high level of evidence for marker based studies.^31^

**Table 1.** Cohorts included in the recurrence and survival pooled analysis by treatment, event and evidence category. All available cohorts which had RSI, physical radiation dose and clinical (recurrence and/or survival) outcomes information were included in the analysis. We denote the number of events, treatment and category of study.^31^ *n.b. A total of 20 patients were censored from the TCGA Glioma cohort and one from the melanoma cohort who were prescribed short course, palliative/hypofractionated radiation doses, as GARD was not designed for hypofractionated regimens.

| Cohort | n | Treatment | Events (rec) | Events (surv) | Category |
| --- | --- | --- | --- | --- | --- |
| Breast (Erasmus)<sup>32,33</sup> | 344 | +RT: 282<br>-RT: 62 | 91<br>12 | -<br>- | C |
| Breast (Karolinska)<sup>34</sup> | 159 | +RT: 77<br>-RT: 82 | 19<br>21 | -<br>- | C |
| Breast, non-TNBC (NKI)<sup>35</sup> | 285 | +RT: 285 | 99 | - | D |
| Breast, TNBC (NKI)<sup>35</sup> | 58 | +RT: 58 | 20 | - | D |
| Endometrium (TCC)<sup>27</sup> | 204 | +RT: 63<br>-RT: 141 | 11<br>7 | 33<br>29 | C |
| Glioma (TCGA)<sup>36</sup> | 244 | +RT: 188<br>-RT: 56 | -<br>- | 134<br>56 | D |
| Head & Neck (NKI)<sup>37</sup> | 92 | +RT: 92 | 28 | - | B |
| Melanoma (TCC)<sup>24</sup> | 41 | +RT: 10<br>-RT: 31 | 5<br>23 | 7<br>22 | C |
| NSCLC (MCC)<sup>38</sup> | 60 | +RT: 60 | 23 | 38 | C |
| Pancreas (TCC)<sup>38</sup> | 73 | +RT: 48<br>-RT: 25 | -<br>- | 33<br>17 | C |
| TNBC (MCC)<sup>38</sup> | 55 | +RT: 55 | 9 | 9 | C |
| Total | 1,615 | +RT: 1218<br>-RT: 397 | 305<br>63 | 254<br>124 |  |

While the range of physical RT dose was limited in these modern cohorts to values near standard of care, delivered in standard fraction sizes, adding the genomic dimension and calculating GARD reveals a wide range of predicted biologic effect (see **Figure 1** and **Supplemental Figures 1**). It should be noted that RSI was derived as a surrogate for the surviving fraction at 2Gy, and therefore our results hold only for doses near to this. All patients in this study were treated with fraction sizes between 1.8 − 2Gy (165 received 1.8Gy, 1 received 1.85Gy and 1, 448 received 2Gy). While only a minor effect, we report all total doses in the form of EQD2 for more appropriate comparison, assuming an *a/b* = 10, though will continue to refer to this as physical, or total RT dose for clarity of exposition. Spatial, intratumoral heterogeneity, while now known as a possible confounder, is not considered here in this large pan-cancer study.

**Figure 1.**
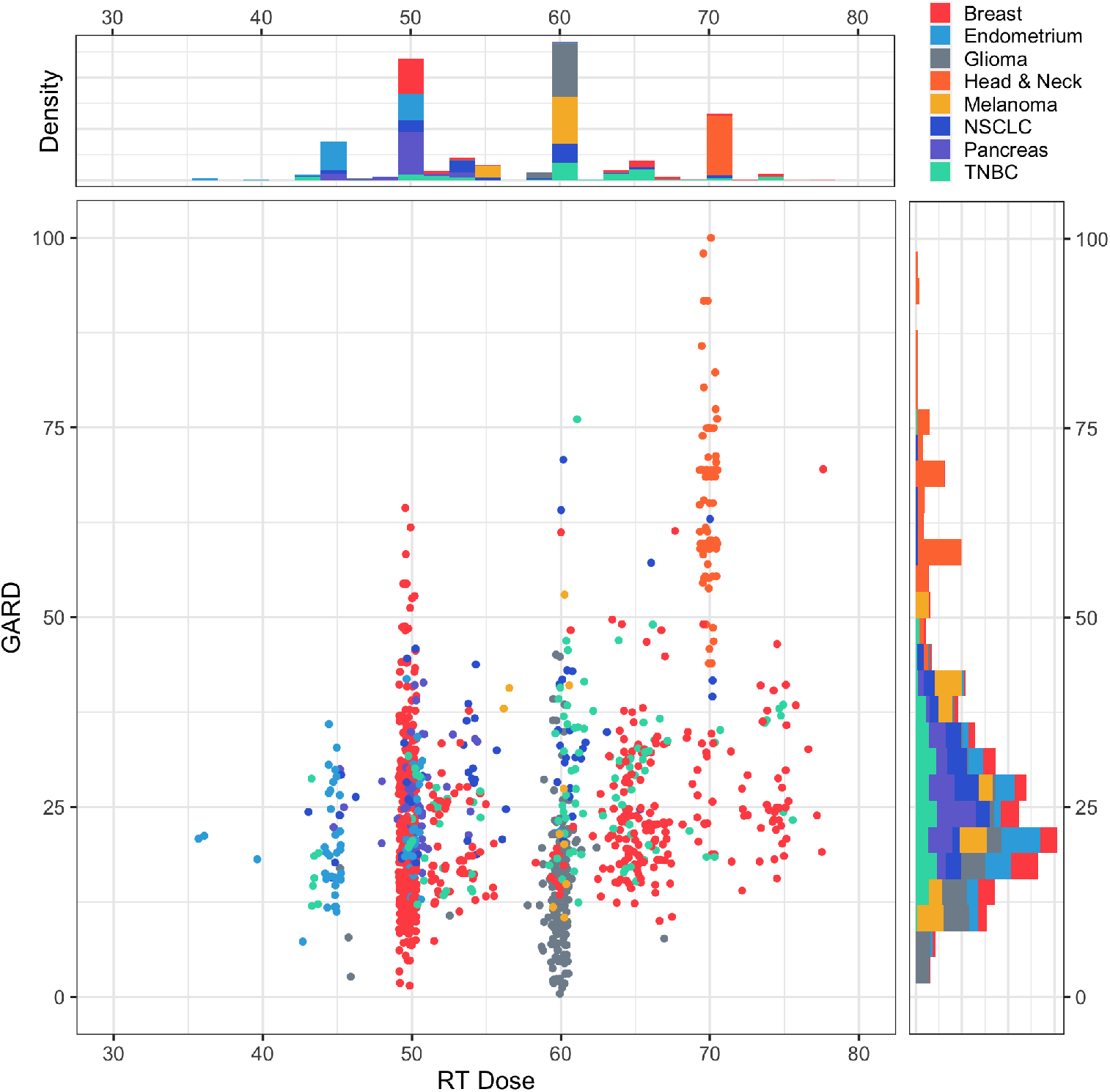
For each patient (colored by disease site) we plot the physical RT dose delivered vs. the associated GARD. While there is a functional relationship between dose and GARD, given the wide underlying genomic heterogeneity, GARD reveals larger ranges, enabling a higher resolution understanding of outcome. For each cohort, the distributions of individual dose and GARD are plotted in stacked histograms on the edges. Several patients with extremely high GARD (*>* 100) are not plotted here for ease of visualization, they can be seen in **Supplemental Figure 1**.

We define two clinical endpoints: time to first recurrence and overall survival. Time to first recurrence included time to local, regional and distant metastasis, and while an imperfect comparator, was forced due to the heterogeneity of the available data, and represented what we think is the most stringent choice. Combining all clinical cohorts, we report the results of two independent analyses of the recurrence and survival endpoints separately. Basic descriptive outcome measures for the clinical cohorts can be seen in **Supplemental Tables 1 and 2**, and Kaplain-Meier plots for each cohort in **Supplemental Figures 2 and 3**. Individual cohorts were analyzed with Cox proportional hazards models. The pooled analysis was analyzed with a Cox proportional hazards model stratified by cohort, and a *c*-squared statistic calculated using the Wald test. Interaction between GARD and whether patients received radiation therapy was calculated for both patient pools using the Wald statistic. Restricted cubic splines with 3 knots (13.1, 23.0, 46.9) were included to allow for possible nonlinear associations.

### Pooled Analysis Method

In order to statistically compare cohorts which are heterogeneous in outcome and disease type, we used a Cox proportional hazards model, stratified by cohort, with GARD as the only covariate. This allowed each cohort to have a different baseline hazard function, yet still determine a common GARD effect. Patients who did not receive RT were pooled together to form a cohort for a similar analysis. To calculate GARD for patients that did not receive RT (sham GARD), we assumed that they all received standard of care dose sham RT (50 Gy in 25 fractions for breast, 50 Gy in 25 fractions for pancreas, 60 Gy in 30 fractions for glioma, 54 Gy in 27 fractions for endometrium). While we realize that this is a strong assumption, this seemed reasonable to determine the association of GARD with recurrence and survival in non-RT treated patients. Of note, this is functionally equivalent to testing for the of RSI as a prognostic biomarker in non-RT treated patients, something we have studied in depth before^21^, as GARD and RSI are rank preserved when all patients receive equivalent physical RT dose. We also utilized restricted cubic splines in order to allow for non linear effects.

### Relationship between distributions of GARD and RT dose within each disease site

The joint distributions of RT dose and GARD reveal the association between the two for each patient, plotted by site in **Figure 1**, with the associated distributions of each plotted as histograms on the edges. In this plot, we combine each cohort by disease type for ease of clinical comparison.

## Results

### GARD is a pan-cancer predictor of RT benefit

To determine the association between GARD and clinical outcome in RT-treated patients, we performed Cox regression analysis for each cohort. The results of this analysis (see **Figure 2, top**) are represented as a forest plot demonstrating the impact of GARD on recurrence and survival endpoints in each of the individual cohorts studied (see **Supplemental Table 3** for this information in tabular form). As a negative control, we performed the same analyses in patients treated without RT (GARD calculated using standard of care dose sham radiation) which showed no association between GARD and either recurrence or overall survival (see **Figure 2, middle** and **Supplemental Table 5**). Pooled analysis of all available data reveal that GARD is associated with recurrence (HR = 0.982, CI [0.970, 0.994], p = 0.002) and survival (HR = 0.970, CI [0.953, 0.988], p = 0.001).

**Figure 2.**
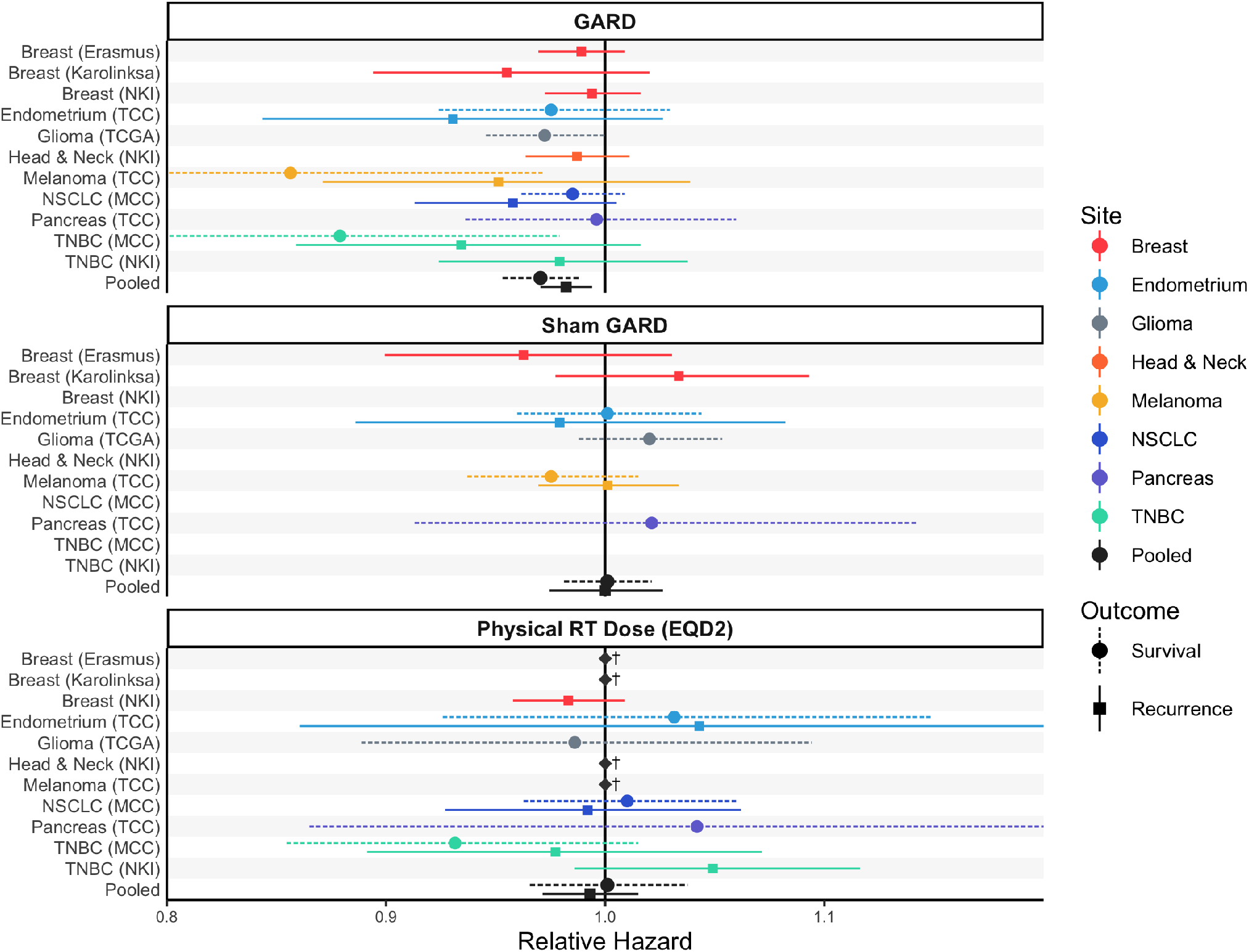
Individual Cox proportional hazard ratios for each disease site and outcome. We present three separate Cox proportional hazards models. **Top:** We use GARD as a covariate for OS and first recurrence for each site individually, and for the composite. In the pooled analysis, GARD is significantly associated with both OS (p = 0.001) and recurrence (p = 0.002). **Middle:** We use sham-GARD as the covariate for the same analysis. These are patients not treated with RT, but we calculated GARD using what would have been standard of care. There are no significant associations. **Bottom:** We use physical RT dose as the covariate for the same analysis; no sites or groups are significantly associated. Recurrence (solid) and survival (dotted) were used as the outcome of interest for all disease sites. Each point represents the hazard ratio with horizontal lines showing 95% CI, which are symmetric (n.b. CIs for Melanoma and TNBC are truncated on the left in the GARD panel, and endometrium and pancreas are truncated on the right in the physical RT dose panel for ease of visual analysis). Values to the left of the intercept, HR = 1, favour a predictive correlation with improved outcome of interest. †: Non-convergence of Cox models secondary to singular dose values.

To determine whether GARD is a pan-cancer predictor of RT benefit, we performed an interaction test between GARD and treatment (with RT or without RT) for both outcomes. The interaction test for survival revealed a significant interaction between GARD and RT treatment status (Wald statistic, p = 0.011, see **Supplemental Table 6**), but not for recurrence (Wald statistic, p = 0.218, see **Supplemental Table 7**). However, using a previously defined cut-point (GARD > 21),^30^ the interaction for recurrence on the linear model is promising (Wald statistic p = 0.060). An exploratory analysis using non linear restricted cubic splines revealed varying associations for different ranges of GARD and recurrence. Although the interaction between RT and GARD for recurrence and survival appears different, this observation is likely related to the different populations included in each analysis (survival and recurrence).

### RT dose does not predict outcome until modulated by genomics (GARD) in RT treated patients

Historically, the RT dosing prescription paradigm has been guided by physical RT dose. We have previously shown that the effect of a given RT dose, as quantified by GARD, varies widely between patients, and is associated with changes in clinical outcome.^6^ To test how GARD compares directly to physical RT dose (EQD2), we performed a second Cox regression analysis for each cohort using physical RT dose as the covariate. We found no association between physical RT dose and survival (p = 0.955) or recurrence (p = 0.532) (**Figure 2, bottom**); in contrast to GARD, which predicted both survival and recurrence in the pooled analysis (see **Supplemental Table 4** for Cox model results in tabular form for physical RT dose).

### Increasing GARD results in a statistically significant decrease in relative hazard for survival and recurrence

Having shown that GARD is predictive of RT benefit, and is superior to physical RT dose, we sought to understand the effect of increasing GARD (increased biological effect delivered, measured in units of GARD) for any patient. We subsequently fit a relative hazard function stratified by disease cohort to assess whether GARD has an association with the outcome of interest (recurrence and survival). The resulting regression found GARD to be a statistically significant, linear variable for both outcome types analyzed, with an increase in GARD corresponding to a decrease in the GARD-specific relative hazard (first recurrence: *c*-squared = 9.9,p = 0.002; overall survival: *c*-squared = 11.5,p = 0.001, see **Supplemental Figure 6**). In order to further convey the clinical significance of these results, disease site and baseline hazard functions were reintroduced in order to predict absolute survival probabilities at 3 years, plotted as a nomogram beneath each outcome model (see **Figure 3**). Several patients (outliers with extremely high GARD) have been removed from the visualization in **Figure 3**, but are included in the analysis/model, and are represented in **Supplemental Figure 6**, and model residuals in **Supplemental Figure 7**.

**Figure 3.**
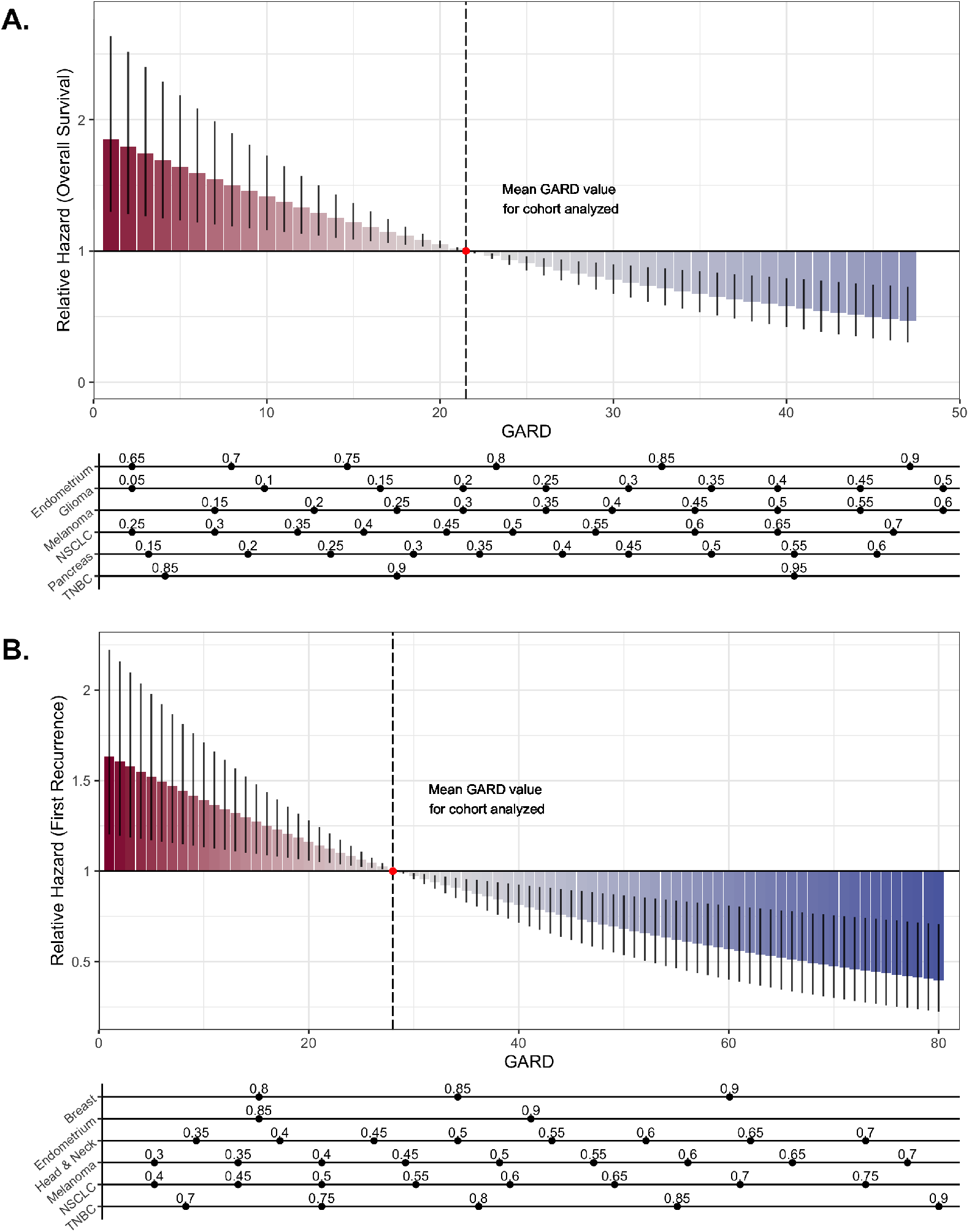
GARD is a statistically significant, continuous, predictive biomarker of survival and recurrence in RT treated patients. We plot the relative hazard per unit GARD at 3-years as predicted by the stratified Cox model for both **(A)** overall survival and **(B)** first recurrence. The impact of the relative hazard as determined by GARD can be interpreted in the context of a specific site, shown as a nomogram below the waterfall, where predicted absolute survival probability underlies the corresponding GARD value for each cohort. n.b. several extremely high GARD outliers have been removed from the plot for ease of visualization, but not from the analysis, and they are included in **Supplemental Figure 6**.

## Discussion

Radiation therapy is the single most commonly utilized anti-cancer therapy and is currently prescribed based on an empiric, one-size-fits-all dosing paradigm. In this analysis we have shown that biological effect, as quantified by GARD, is statistically associated with improvements in recurrence and overall survival in RT treated patients. Critically, we demonstrate that the interaction of GARD and RT is significant for overall survival, establishing GARD as predictive of RT-treatment benefit. We utilized all available clinical data, consisting of 11 cohorts, totalling 1,615 patients spanning 7 disease sites. In a recent commentary the EORTC summarized the clinical data supporting RSI/GARD as representing “near level one evidence” for its utilization. Furthermore, Khan and colleagues recently published validation of RSI in bladder cancer samples from a recent Phase 3 prospective clinical trial (Category B study). ^39^ Although in Simon’s classification of biomarker studies he did not specify a role for meta-analyses, we propose that the combined evidence developed for RSI/GARD represents level one evidence (consistent results in multiple category b and c studies including this meta-analysis). However, we acknowledge that definitive evidence would come from a randomized clinical trial.

Like all pooled/meta-analyses, ours suffers from some limitations, mostly secondary to the heterogeneous nature of the cohorts and interventions. In particular, comparing patients across disease sites makes more standard clinico-pathologic comparisons impossible given the differences in these variables between disease sites, and there is a subsequent lack of overlap in the cohorts. Full analyses were completed in each individual cohort in previous publications, showing the independent value of RSI/GARD controlling for other (disease specific) variables, including chemotherapy. Additionally, given the current limitation of RSI/GARD to understand dosing near standard fractionation, we are unable to properly analyze or model some of the newer hypo-or hyper-fractionated regimens, and given the heterogeneity of our cohorts, are unable to address chemoradiation specific questions here. These limitations aside, a paradigm driven by GARD defines, for the first time, an actionable measure of biological effect of RT in a given patient. This permits a reframing of our thinking, enabling us to modulate and personalize the biological effect of radiotherapy we deliver to our patients, not just the physical dose, and establishes the first clinically-validated approach to genomic-based radiation oncology.

The results of our pan-cancer pooled analysis demonstrate that the biologic effect induced by physical RT dose, as quantified by GARD, is associated with both survival and recurrence, while the physical RT dose is not. This makes sense in the setting of recent clinical trials of RT dose escalation in unselected patients, which have not been shown to improve overall survival – and does not mean that physical RT dose is not an important parameter. Because the biological effect of a given dose varies widely for individual patients, small changes near the ranges now accepted as standard of care are not uniform, so the effects can be obfuscated in unselected patients. This suggests that to make further progress, like in medical oncology, we need to incorporate the extra dimension of genomics to provide a higher resolution understanding of the biological effect of the RT dose we are delivering to our patients and and how this can affect their outcomes. Thus, it is critical to shift the radiotherapy dosing paradigm from one that is based on physical RT dose to one that is based on biological effect, as quantified by GARD. The fundamental goal of RT planning is to deliver the prescribed physical RT dose to the target volume while minimizing dose to normal tissue. The integration of 3D anatomy into radiation treatment planning systems, brought about by the invention of the CT scanner, enabled the geometric optimization of radiation fields. This led to a shift from one-size-fits-all radiotherapy field shapes to ones that are anatomically personalized, improving the ability of radiation oncologists to further spare normal tissue for each individual patient. The resultant techniques (e.g. Intensity Modulated Radiation Therapy (IMRT), Stereotactic Body Radiation Therapy (SBRT)) have been shown to decrease toxicity in Phase 3 clinical trials and change standard of care in several cancers including lung and prostate. In other words, the development of radiation planning optimization techniques led to significant clinical gains for RT-treated patients. Similarly, we propose that GARD can be utilized to maximize the biological effect of physical dose to the tumor, while respecting standard of care guidelines for normal tissue. In a recent study, we demonstrated that GARD-based optimization of tumor physical dose can be achieved within standard of care guidelines for up to 75% of non-small cell lung cancer patients treated with post-operative RT.^40^ In this work, we show that GARD can be used to rationally optimize (through escalation or de-escalation) radiation benefit and toxicity, something that has not been possible with physical RT dose alone. As the first genomic framework to predict RT benefit, we believe the clinical utility of GARD will be paramount. GARD is the first model to provide the clinical radiation oncologist with decision support information to modulate the potential RT benefit for each individual tumor. In addition, it quantifies the effect size for each individual patient. This will provide radiation oncologists with a complete set of new tools to optimize RT dose using genomics. Importantly, GARD has been established in the CLIA laboratory at Moffitt Cancer Center, and clinical trials are expected to start accruing later this year. Finally, we are not suggesting abandoning physical RT dose, but instead, as the CT scanner did for x-ray, we suggest enhancing dose with another dimension, genomic data, allowing us to see each individual patient’s potential for RT benefit at a higher resolution.

Recently, multiple phase 3 randomized clinical trials failed to demonstrate improvements in clinical outcome after uniform dose escalation. How do you reconcile the results of these trials with the proposal that each unit increase in GARD positively impacts clinical outcome? While this stands in contrast to the recent negative trials of dose-escalation, it is not dissonant. First, given our new understanding of heterogeneous response to homogeneous physical RT dose, we realize that uniform dose escalation without understanding intrinsic radiosensitivity is actually not uniform. A GARD-based approach allows for an understanding of this non-uniform response to dose escalation – some patients benefit while others are potentially harmed they receive unnecessary extra dose and associated toxicity – such as is the leading hypothesis for the results of RTOG0617.^41^ Instead of rejecting the hypothesis that dose-escalation is beneficial, GARD allows us see which patients would benefit most from dose optimization – permitting next generation trials of personalized RT dose.

Today, radiation therapy is prescribed using an empirically derived, one-size-fits-all approach, a paradigm in which patients with a given cancer type receive uniform doses of RT. This pooled analysis demonstrates that GARD, a quantification of the biological effect of RT dose, is predictive of RT treatment benefit and our standard measure of radiation dose is not. We postulate that a GARD-based framework of RT should be adopted as the new paradigm for trial design, radiation dosing and personalized medicine in radiation oncology.

## Data Availability

All data are published previously in this pooled analysis. The code (and data) are available on github.

https://github.com/gsedor/GARD_Meta-Analysis

