## Supplemental Material for "GARD is a pan-cancer predictor of radiation therapy benefit"

### Supplemental Information

#### Code and data availability

All code and data for the analyses in this paper can be found on Github: github.com/gsedor/GARD_ Meta-Analysis.

#### Calculation of GARD

As previously described,[6] GARD is derived using the linear-quadratic model ($S = e^{-nd(\alpha+\beta d)}$), and the individual RSI and radiation dose/fractionation schedule for each patient. RSI was previously trained in 48 cancer cell lines to predict cellular radiosensitivity as determined by survival fraction at 2 Gy (SF2.)[16] Each of ten genes in the algorithm is ranked based on gene expression (highest expressed gene is ranked at 10 and lowest at 1) and RSI is calculated using the pre-determined equation:

$$\begin{aligned} \mathrm{RSI} = -0.0098009 \times \mathrm{AR} + 0.0128283 \times \mathrm{cJun} + 0.0254552 \times \mathrm{STAT1} - 0.0017589 \times \mathrm{PKC} - \\ 0.0038171 \times \mathrm{RelA} + 0.1070213 \times \mathrm{cABL} - 0.0002509 \times \mathrm{SUMO1} - \\ 0.0092431 \times \mathrm{PAK2} - 0.0204469 \times \mathrm{HDAC1} - 0.0441683 \times \mathrm{IRF1}. \end{aligned} \tag{1}$$

To obtain GARD, first, a patient-specific (genomic) $\alpha_g$ is derived by substituting RSI for Survival (S) in the classic LQ equation, yielding:

$$\alpha_g = -\frac{\ln \mathrm{RSI}}{nd} - \beta d, \tag{2}$$

where dose ($d$) is 2Gy, $n$ is the number of fractions (here $n = 1$), and $\beta$ is a constant $0.05/\mathrm{Gy}^2$, an assumption we plan to study in a disease site by disease site manner in future work. GARD is then calculated using the equation for biologic effect: $\mathrm{GARD} = nd(\alpha_g + \beta d)$, the patient-specific $\alpha_g$ calculated as per equation (2), and the number of fractions ($n$) and dose per fraction ($d$) received by each patient per the reported trials. Joint and individual distributions for dose, RSI and GARD for each patient in this analysis can be seen in **Figure 1** and **Supplemental Figures 1** respectively.

#### Treatment and outcome measures for each included cohort

##### *Description of each individual cohort*

For each of the cohorts analyzed, we provide basic descriptive treatment and outcome measures. For the recurrence analysis, the pooled cohort includes a total of 1,298 (982 +RT, 316 -RT) patients to analyze, with 368 events (305 +RT, 63 -RT). The survival analysis included 677 patients (424 +RT, 253 -RT) with 378 events (254 +RT, 124 -RT). Recurrence pool for patients treated with RT (+RT) consisted of 982 patients at risk in 9 separate cohorts with 305 events, and survival pool consisted of 424 patients at risk in 6 cohorts with 254 events. Recurrence pool for patients not treated with RT (sham-GARD) consisted of 316 patients at risk in 4 separate cohorts with 63 events, and survival pool consisted of 253 patients at risk in 4 cohorts with 124 events.

Here, we will provide a more granular description of each included cohort, including details on how the genomic data was obtained and whether the data was part of a prospective trial. We rate the category of study as proposed by Simon in his work "Use of Archived Specimens in Evaluation of Prognostic and Predictive Biomarkers," which defines the category of the level of evidence for an individual cohort.[31] This is important as the dichotomy between prospective and retrospective is less important for archival tissue studies when determining the level of conclusions once can draw.

1. Breast Cancer - Erasmus Cohort[32,33]: This cohort includes 344 lymph node negative breast cancer patients treated without systemic chemotherapy. All clinical details were previously published. Samples were obtained from the NKI reference laboratory where samples were routinely submitted. All clinical data was routinely collected according to a standard protocol. Clinical analysis was retrospective. Based on the Simon criteria this is a Category C study.

2. Breast Cancer - Karolinska Cohort[34]: This study is part of a prospective/observational study and clinical details have been previously described. It includes a total 159 patients. Follow up data was prospectively collected by the Swedish

Tumor Registry. Clinical decisions were not dictated by protocol. 104 patients received adjuvant CMF. Clinical analysis was retrospective. Based on the Simon's criteria this is a Category C study.

3. Breast Cancer - NKI, Institut Curie Cohort (non-TNBC)[35]: This cohort includes 285 patients from four Dutch institutions and the Institut Curie All clinical details have been extensively described. It was specifically developed to identify a signature of local recurrence after breast conservation therapy. All clinical and genomic analysis were retrospective. Clinical treatment was not protocol-dictated. Based on Simon's criteria this is a Category D study.

4. Breast Cancer - NKI, Institut Curie Cohort (TNBC)[35]: This cohort includes 58 patients from four Dutch institutions and the Institut Curie All clinical details have been extensively described. It was specifically developed to identify a signature of local recurrence after breast conservation therapy. All clinical and genomic analysis were retrospective. Clinical treatment was not protocol-dictated. Based on Simon's criteria this is a Category D study.

5. Endometrial cancer - Moffitt Cancer Center: This cohort included 204 endometrial cancer patients. Clinical details were previously published. Clinical treatment was not protocol-dictated. Genomic profiling and clinical outcome was done as part of a prospective/observational tissue collection protocol at Moffitt (Total Cancer Care). Outcome data was confirmed with chart review. Based on the Simon criteria this is a Category C study.

6. Glioblastoma - TCGA[36]: This cohort includes a total of 270 GBM patients. Clinical details were previously published. Genomic profiling was done as part of TCGA and were all retrospectively performed. Clinical treatment was not protocol-dictated. Based on the Simon's criteria this is a Category D study.

7. Head and neck cancer - NKI Cohort[37]: This cohort includes 92 locally-advanced head and neck cancer patients treated with concurrent chemoradiation within prospective Phase 2-3 clinical trials at the Netherlands Cancer Institute (NKI). Clinical details were previously reported. Treatment and follow up were prospective and protocol dictated. Genomic analysis was retrospective but tissue was collected prospectively. Based on the Simon's criteria this is a Category B study.

8. Melanoma - Moffitt Cancer Center[23]: This cohort includes 42 patients that were prospectively genomically profiled as part of a prospective/observation tissue collection protocol at Moffitt Cancer Center (Total Cancer Center). The clinical details of the cohort have been previously published. Clinical treatment was not protocol-dictated. Genomic profiling and clinical outcome was done as part of a prospective/observational tissue collection protocol at Moffitt (Total Cancer Care). Outcome data was confirmed with chart review. Based on the Simon's criteria this is a Category C study.

9. Lung cancer - Moffitt Cancer Center: This cohort includes 60 patients with Non-small cell lung cancer (NSCLC) treated with post-operative RT. Clinical details have been previously published. Clinical treatment was not protocol-dictated. Genomic profiling and clinical outcome was done as part of a prospective/observational tissue collection protocol at Moffitt (Total Cancer Care). Outcome data was confirmed with chart review. Based on the Simon's criteria this is a Category C study.

10. Pancreas cancer - Mofitt Cancer Center: This cohort included 73 patients and clinical details have been previously described. Clinical treatment was not protocol-dictated. Genomic profiling and clinical outcome was done as part of a prospective/observational tissue collection protocol at Moffitt (Total Cancer Care). Outcome data was confirmed with chart review. Based on the Simon criteria this is a Category C study.

11. Triple negative breast cancer - Moffitt Cancer Center[38]: This cohort includes 55 triple negative breast cancer patients that were prospectively genomically-profiled as part of a prospective-observational study at Moffitt Cancer Center (Total Cancer Care). The clinical details have been previously published. The genomic analysis was prospectively performed following a pre-defined protocol for tissue processing and gene expression. Clinical data was prospectively obtained but follow up and treatment were not dictated by protocol and followed clinical standard of care as determined by the patient's physician. Based on the Simon's criteria this is a Category C study.

##### ***Physical RT dose and GARD distributions***

Physical RT dose by itself lacks the resolution to determine changes in outcome at the individual patient level, and necessarily has limited ranges in the era of evidence-based medicine when we are limited to trials of uniform dose-escalation. Adding a genomic dimension to dose, using GARD, reveals a wide heterogeneity in predicted biologic effect of even doses of limited (or no) range. We plot the distributions of physical RT dose, and associated GARD, in **Supplemental Figure 1**.

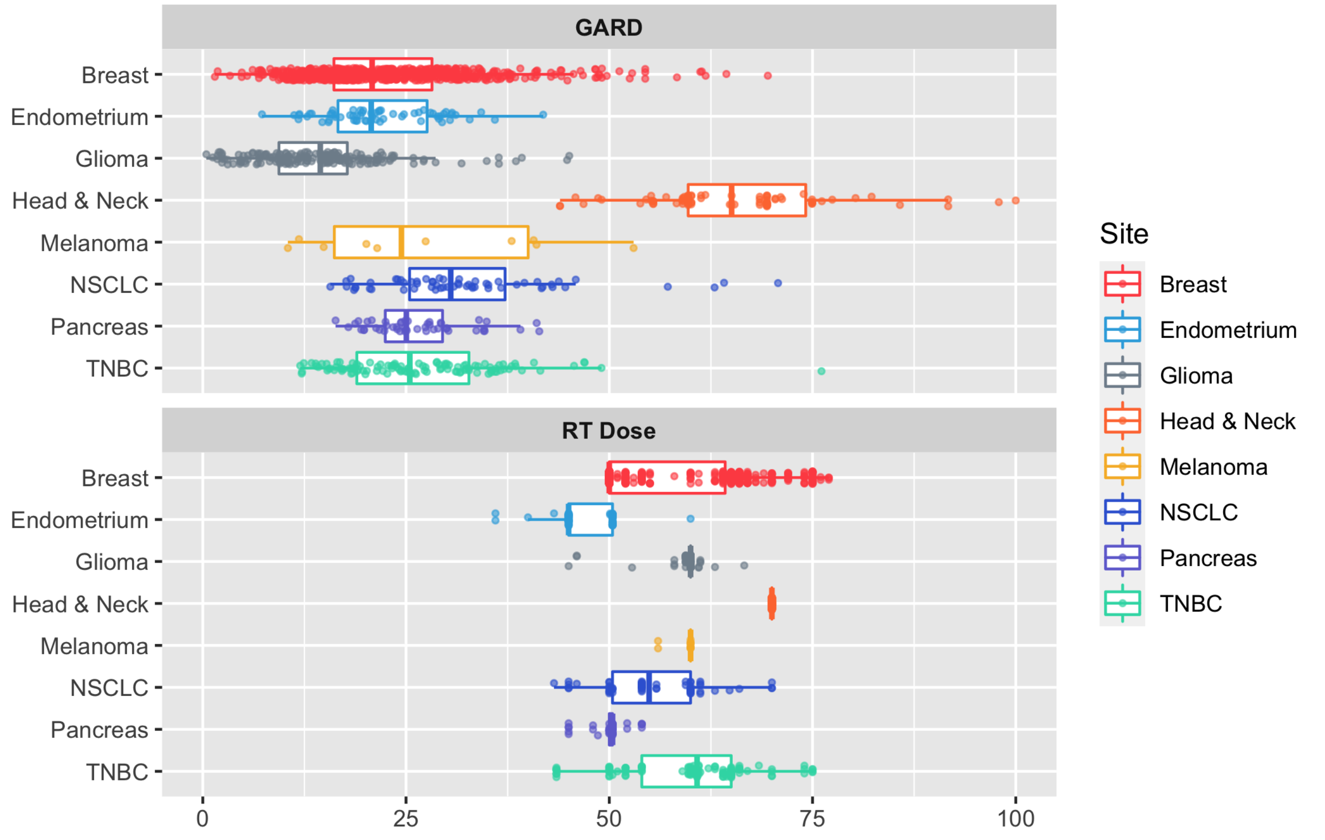


**Supplemental Figure 1. For each cohort we plot the physical RT dose delivered and the associated GARD.** The range of physical RT dose is quite limited, and near to the standard of care for each cohort as expected, making it difficult to associate differences in outcome related to dose. Adding a genomic dimension to dose, using GARD, reveals wide heterogeneity within each group invisible before.

##### *Recurrence cohorts*

In **Supplemental Table 1** we present the 1- and 5-year proportions of patients without recurrence for each cohort, with and without RT, as applicable (if no patients exist in a given cohort, it is labeled NA).

**Supplemental Table 1. Proportion of patients without recurrence at 1- and 5-years, +/- RT.**

| Cohort | 1-yr (-RT) | 1-yr (+RT) | 5-yr (-RT) | 5-yr (+RT) |
|---|---|---|---|---|
| Erasmus_breast[32, 33] | 0.952 [0.900, 1.000] | 0.936 [0.908, 0.965] | 0.806 [0.714, 0.911] | 0.677 [0.624, 0.733] |
| Karolinksa_breast[34] | 0.950 [0.904, 0.999] | 0.987 [0.962, 1.000] | 0.747 [0.657, 0.849] | 0.805 [0.721, 0.899] |
| MCC_lung[38] | NA | 0.809 [0.709, 0.924] | NA | 0.534 [0.400, 0.714] |
| MCC_breast_TN[38] | NA | 0.944 [0.885, 1.000] | NA | 0.844 [0.749, 0.950] |
| NKI_breast[35] | NA | 1.000 [1.000, 1.000] | NA | 0.740 [0.691, 0.793] |
| NKI_breast_TNBC[35] | NA | 0.862 [0.778, 0.956] | NA | 0.672 [0.562, 0.805] |
| Pranama_HN[37] | NA | 0.741 [0.653, 0.842] | NA | 0.618 [0.486, 0.787] |
| TCC_endometrial[27] | 0.963 [0.929, 0.999] | 0.968 [0.925, 1.000] | 0.924 [0.869, 0.982] | 0.768 [0.646, 0.914] |
| TCC_melanoma[24] | 0.710 [0.567, 0.889] | 0.700 [0.467, 1.000] | 0.238 [0.121, 0.468] | 0.450 [0.211, 0.961] |

In **Supplemental Figure 2** we present the associated Kaplan-Meier curves for first recurrence for all cohorts analyzed.

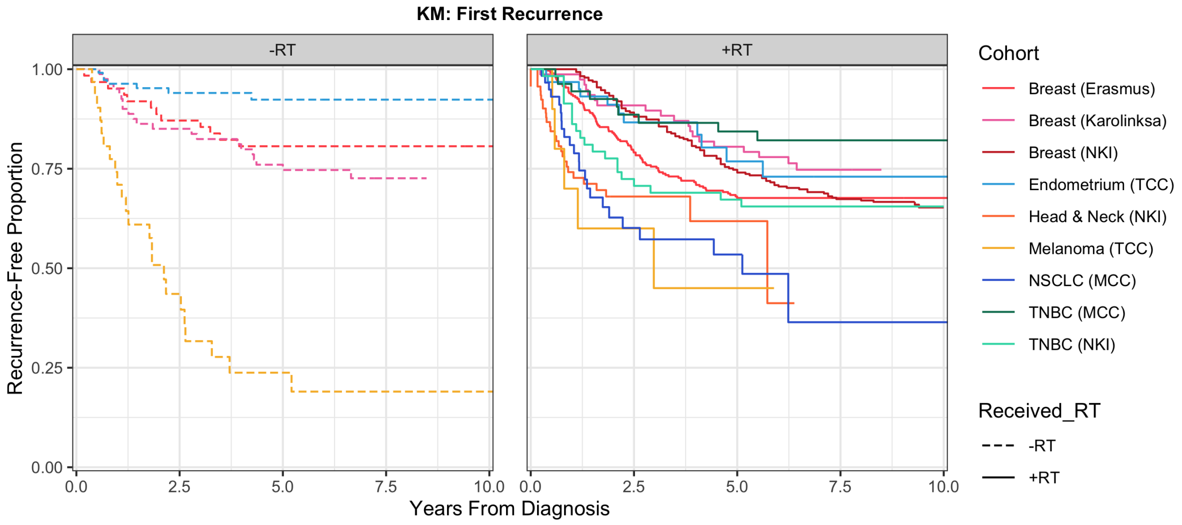


**Supplemental Figure 2. Kaplan-Meier curves for each cohort, both without (left) and with (right) RT with first recurrence as the outcome.**

##### *Overall survival cohorts*

In **Supplemental Table 2** we present the 1- and 5-year overall survival for each cohort, with and without RT, as applicable (if no patients exist in a given cohort, it is labeled NA).

**Supplemental Table 2. Proportion of patients surviving at 1- and 5-years, with and without radiation treatment**

| Cohort | 1-yr (-RT) | 1-yr (+RT) | 5-yr (-RT) | 5-yr (+RT) |
|---|---|---|---|---|
| MCC_lung[38] | 0.883 [0.806, 0.968] | NA | 0.440 [0.327, 0.592] | NA |
| MCC_TNBC[38] | 1.000 [1.000, 1.000] | NA | 0.882 [0.797, 0.975] | NA |
| TCC_endometrial[27] | 0.935 [0.875, 0.999] | 0.967 [0.936, 0.999] | 0.531 [0.411, 0.686] | 0.774 [0.691, 0.866] |
| TCC_melanoma[24] | 0.900 [0.732, 1.000] | 0.903 [0.805, 1.000] | 0.300 [0.116, 0.773] | 0.388 [0.243, 0.619] |
| TCC_pancreas[38] | 0.771 [0.661, 0.899] | 0.800 [0.658, 0.973] | 0.352 [0.239, 0.517] | 0.279 [0.142, 0.550] |
| TCGA_glioma[17] | 0.736 [0.670, 0.808] | 0.054 [0.018, 0.161] | 0.056 [0.026, 0.120] | NA |

In **Supplemental Figure 3** we present the associated Kaplan-Meier curves for OS for all cohorts analyzed.

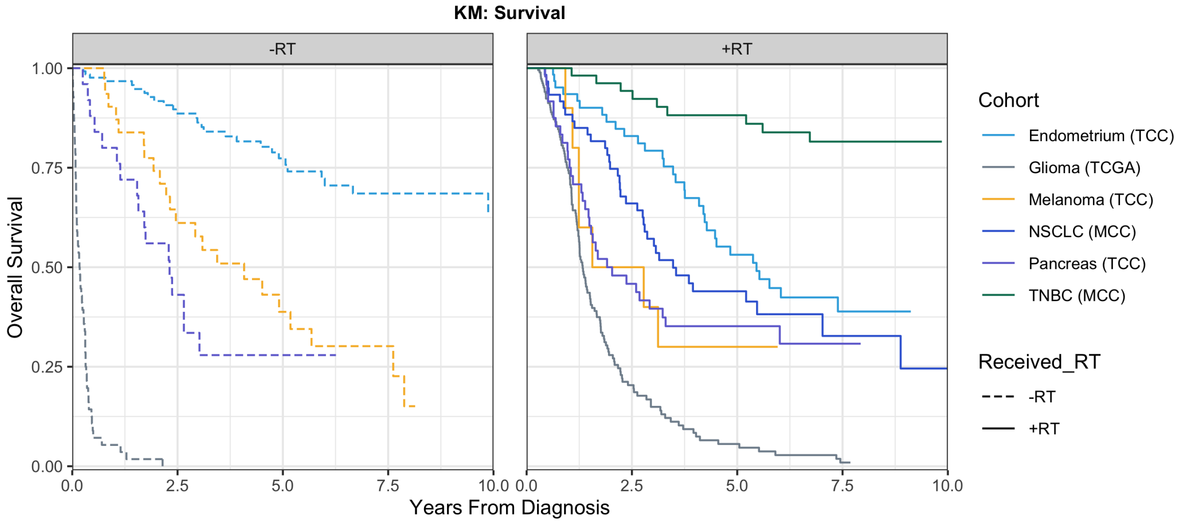


**Supplemental Figure 3. Kaplan-Meier curves for each cohort, both without (left) and with (right) RT with overall survival as the outcome.**

#### Individual cohort and pooled Cox model results

We present the results of the individual Cox models for GARD, physical RT dose and sham-GARD as presented in the forest plot in **Figure 2** in tabular form.

##### *GARD*

GARD is significantly associated with both overall survival and recurrence in the pooled analysis (see **Supplemental Table 3**).

| | Recurrence | | | | | | Survival | | | | | |
|---|---|---|---|---|---|---|---|---|---|---|---|---|
| | | | HR | | | | | | HR | | | |
| Site | n | event | est | lower | upper | p | n | event | est | lower | upper | p |
| Breast (Erasmus)[32,33] | 282 | 91 | 0.989 | 0.969 | 1.009 | 0.261 | NA | NA | NA | NA | NA | NA |
| Breast (Karolinksa)[34] | 77 | 19 | 0.955 | 0.894 | 1.020 | 0.161 | NA | NA | NA | NA | NA | NA |
| Breast (NKI)[35] | 285 | 99 | 0.994 | 0.972 | 1.016 | 0.582 | NA | NA | NA | NA | NA | NA |
| Endometrium (TCC)[27] | 63 | 11 | 0.931 | 0.844 | 1.026 | 0.139 | 63 | 33 | 0.975 | 0.924 | 1.029 | 0.353 |
| Glioma (TCGA)[36] | NA | NA | NA | NA | NA | NA | 188 | 134 | 0.972 | 0.946 | 1.000 | 0.042 |
| Head & Neck (NKI)[37] | 92 | 28 | 0.987 | 0.964 | 1.011 | 0.269 | NA | NA | NA | NA | NA | NA |
| Melanoma (TCC)[23] | 10 | 5 | 0.951 | 0.871 | 1.039 | 0.248 | 10 | 7 | 0.856 | 0.755 | 0.971 | 0.013 |
| NSCLC (MCC)[38] | 60 | 23 | 0.958 | 0.913 | 1.005 | 0.069 | 60 | 38 | 0.985 | 0.962 | 1.009 | 0.228 |
| Pancreas (TCC)[38] | NA | NA | NA | NA | NA | NA | 48 | 33 | 0.996 | 0.936 | 1.060 | 0.887 |
| TNBC (MCC)[38] | 55 | 9 | 0.934 | 0.859 | 1.016 | 0.106 | 55 | 9 | 0.879 | 0.789 | 0.979 | 0.017 |
| TNBC (NKI)[35] | 58 | 20 | 0.979 | 0.924 | 1.038 | 0.467 | NA | NA | NA | NA | NA | NA |
| Pooled | 982 | 305 | 0.982 | 0.970 | 0.994 | **0.002** | 424 | 254 | 0.970 | 0.953 | 0.988 | **0.001** |

**Supplemental Table 3. Hazard ratios for each disease site and outcome type with GARD as the covariate.** Each cohort was fit to a unique Cox proportional hazard function using GARD as the covariate in order to quantify the impact of each cohort on the final analysis. GARD is significantly associated with both overall survival and recurrence in the pooled analysis.

##### *Physical RT Dose*

Physical RT dose (**Supplemental Table 4**) has no statistically significant associations with any outcome.

| | Recurrence | | | | | | Survival | | | | | |
|---|---|---|---|---|---|---|---|---|---|---|---|---|
| | | | HR | | | | | | HR | | | |
| Site | n | event | est | lower | upper | p | n | event | est | lower | upper | p |
| Breast (Erasmus)[32,33] | † | † | † | † | † | † | † | † | † | † | † | † |
| Breast (Karolinksa)[34] | † | † | † | † | † | † | † | † | † | † | † | † |
| Breast, non-TNBC (NKI)[35] | 285 | 99 | 0.983 | 0.958 | 1.009 | 0.182 | NA | NA | NA | NA | NA | NA |
| Endometrium (TCC)[27] | 63 | 11 | 1.043 | 0.861 | 1.264 | 0.663 | 63 | 33 | 1.031 | 0.926 | 1.149 | 0.561 |
| Glioma (TCGA)[36] | NA | NA | NA | NA | NA | NA | 188 | 134 | 0.986 | 0.889 | 1.094 | 0.782 |
| Head & Neck (NKI)[37] | † | † | † | † | † | † | † | † | † | † | † | † |
| Melanoma (TCC)[23] | 10 | 5 | 8.793 | 0.000 | $1.22e12$ | 0.865 | 10 | 7 | 8.846 | 0.000 | $1.22e10$ | 0.836 |
| NSCLC (MCC)[38] | 60 | 23 | 0.992 | 0.927 | 1.062 | 0.817 | 60 | 38 | 1.010 | 0.963 | 1.060 | 0.686 |
| Pancreas (TCC)[38] | NA | NA | NA | NA | NA | NA | 48 | 33 | 1.042 | 0.865 | 1.255 | 0.657 |
| TNBC (MCC)[38] | 55 | 9 | 0.977 | 0.891 | 1.071 | 0.626 | 55 | 9 | 0.931 | 0.855 | 1.015 | 0.101 |
| Breast, TNBC (NKI)[35] | 58 | 20 | 1.049 | 0.986 | 1.116 | 0.120 | NA | NA | NA | NA | NA | NA |
| Pooled | 982 | 305 | 0.993 | 0.971 | 1.015 | **0.532** | 424 | 254 | 1.001 | 0.966 | 1.038 | **0.955** |

**Supplemental Table 4. Hazard ratios for each disease site and outcome type with physical RT dose as the covariate.** Each cohort was fit to a unique Cox proportional hazard function using physical RT dose as the covariate in order to quantify the impact of each cohort on the final analysis. Physical RT dose is not associated statistical with outcome of any kind in the individual cohorts, or the pooled analysis. †: All patients were given the same physical RT dose, so a Cox model could not be fit.

###### *Sham-GARD*

Sham-GARD was calculated for patients not receiving RT as a negative control using what would be the standard of care RT dose for a given disease. Sham-GARD (**Supplemental Table 5**) has no statistically significant associations with any outcome.

| | **Recurrence** | | | | | | **Survival** | | | | | |
|---|---|---|---|---|---|---|---|---|---|---|---|---|
| | | | HR | | | | | | HR | | | |
| Site | n | event | est | lower | upper | p | n | event | est | lower | upper | p |
| Breast (Erasmus)[32,33] | 62 | 12 | 0.963 | 0.899 | 1.030 | 0.258 | NA | NA | NA | NA | NA | NA |
| Breast (Karolinksa)[34] | 82 | 21 | 1.034 | 0.977 | 1.093 | 0.236 | NA | NA | NA | NA | NA | NA |
| Endometrium (TCC)[27] | 141 | 7 | 0.979 | 0.886 | 1.082 | 0.679 | 141 | 29 | 1.001 | 0.960 | 1.044 | 0.980 |
| Glioma (TCGA)[36] | NA | NA | NA | NA | NA | NA | 56 | 56 | 1.018 | 0.982 | 1.055 | 0.307 |
| Melanoma (TCC)[23] | 31 | 23 | 1.001 | 0.969 | 1.034 | 0.934 | 31 | 22 | 0.975 | 0.937 | 1.015 | 0.198 |
| Pancreas (TCC)[38] | NA | NA | NA | NA | NA | NA | 25 | 17 | 1.021 | 0.913 | 1.142 | 0.713 |
| Pooled | 316 | 63 | 1.000 | 0.974 | 1.026 | **0.999** | 253 | 124 | 0.998 | 0.976 | 1.020 | **0.868** |

**Supplemental Table 5. Hazard ratios for each disease site and outcome type with sham-GARD as the covariate.** Each cohort was fit to a unique Cox proportional hazard function using sham-GARD as the covariate in order to quantify the impact of each cohort on the final analysis. We calculated sham-GARD as we would for GARD, but as these patients did not receive RT, we used the standard of care for each disease. Sham-GARD is not associated statistical with outcome of any kind in the individual cohorts, or the pooled analysis.

#### Interaction Tests

To test if GARD is predictive of patient outcome in RT treated patients we performed an interaction test using the Wald statistic between GARD and whether or not a patient received RT.

##### *GARD is predictive of survival in RT treated patients*

GARD is associated with OS in the entire cohort (p = 0.012) and the interaction GARD * RT is statistically significant (**Supplementary Figure 4** and **Supplementary Table 6**, p= 0.011).

**Supplemental Table 6. Wald statistic to test significance for Radiation and GARD in survival cohort**

| | $\chi$-Square | d.f. | p |
|---|---|---|---|
| Received_RT | 38.252 | 2.000 | <0.001 |
| GARD | 8.836 | 2.000 | 0.012 |
| Received_RT * GARD | 6.502 | 1.000 | **0.011** |
| TOTAL | 42.749 | 3.000 | <0.001 |

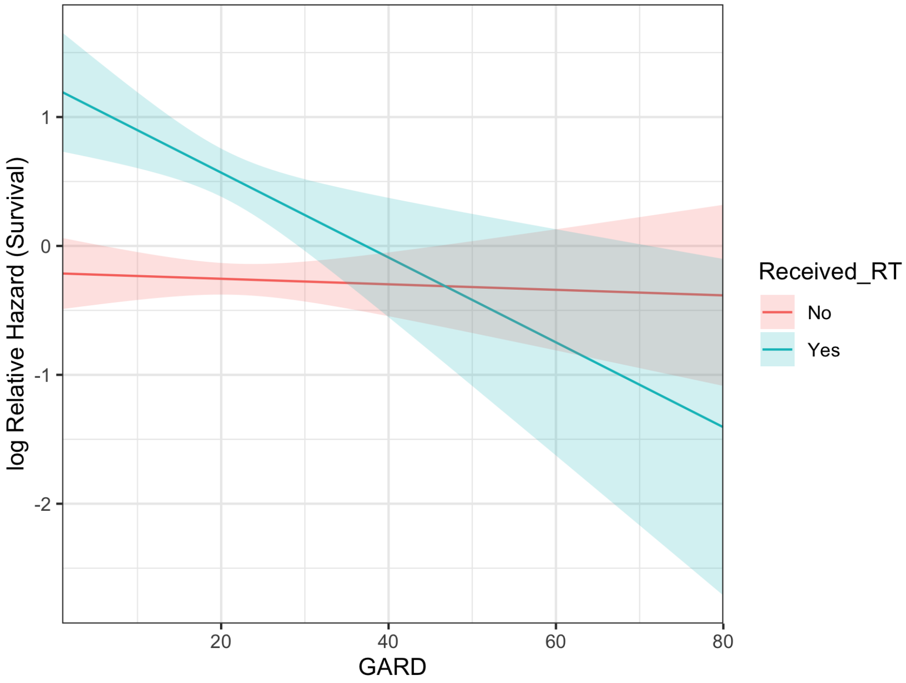


**Supplemental Figure 4. The log Relative Hazard for GARD and overall survival, for those who did and did not receive RT, illustrating the interaction between RT and GARD.**

***GARD is promising as predictive of recurrence in RT treated patients with GARD*** $> 21$***Gy***

GARD is associated with recurrence in the entire cohort (p = 0.005) but the interaction RT*GARD for recurrence is not statistically significant (**Supplemental Table 7**, p = 0.218).

**Supplemental Table 7. Wald statistic to test significance for Radiation and GARD in recurrence cohort**

| | $\chi$-Square | d.f. | p |
|---|---|---|---|
| Received_RT | 6.174 | 2.000 | 0.046 |
| GARD | 10.456 | 2.000 | 0.005 |
| Received_RT * GARD | 1.518 | 1.000 | **0.218** |
| TOTAL | 14.480 | 3.000 | 0.002 |

When patients with extremely low GARD are removed (GARD $<$ 21Gy, based on previously published cutoffs,[30] see **Supplemental Figure 5**), GARD is promising as predictive for first recurrence for patients who received RT (**Supplemental Table 8**, p = 0.060). This is most likely due to the small signal in the group of patients who received the lowest radiation effect.

**Supplemental Table 8. Wald statistic to test significance for Radiation and GARD in recurrence cohort (GARD>21)**

| | $\chi$-Square | d.f. | p |
|---|---|---|---|
| Received_RT | 4.022 | 2.000 | 0.134 |
| GARD | 4.674 | 2.000 | 0.097 |
| Received_RT * GARD | 3.530 | 1.000 | **0.060** |
| TOTAL | 5.130 | 3.000 | 0.163 |

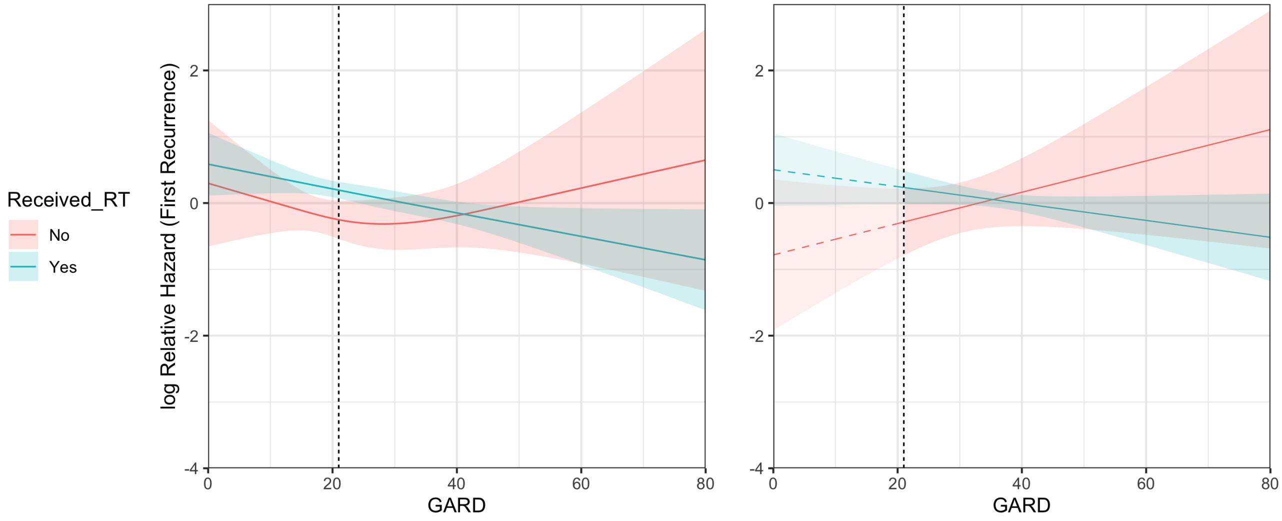


**Supplemental Figure 5. The log Relative Hazard for GARD and recurrence, for those who did and did not receive RT.** The regression was fit using a restricted cubic splines function to assess regions in which GARD interacts with RT with knot locations at 13.1, 23.0, and 46.9 (**left**). A linear model was then used to analyze the subset of patients that had a GARD > 21, (**right**), based on a previously published threshold.

##### Stratified Regression Results

###### *Recurrence and Survival with and without RT*

We present the stratified Cox models for first recurrence and overall survival vs GARD for both cohorts (treated with and without radiation) as a linear model (n.b. these are the same results presented in **Figure 3**, just without the nomogram and in a linear analyses). GARD is continuously associated with first recurrence for radiation treated patients (coefficient $= -0.018$, $\chi$-squared $= 9.9$, $p = 0.002$), and is not for patients not treated with radiation (sham-GARD, coefficient $< 0.001$, $\chi$-squared $< 0.001$, $p = 0.999$), see **Supplemental Figure 6, left**). GARD is a continuous predictor of overall survival for radiation treated patients (coefficient $= -0.030$, $\chi$-squared $= 11.5$, $p = 0.001$), and is not for patients not treated with radiation (sham-GARD, coefficient $= -0.002$, $\chi$-squared $< 0.01$, $p = 0.868$).

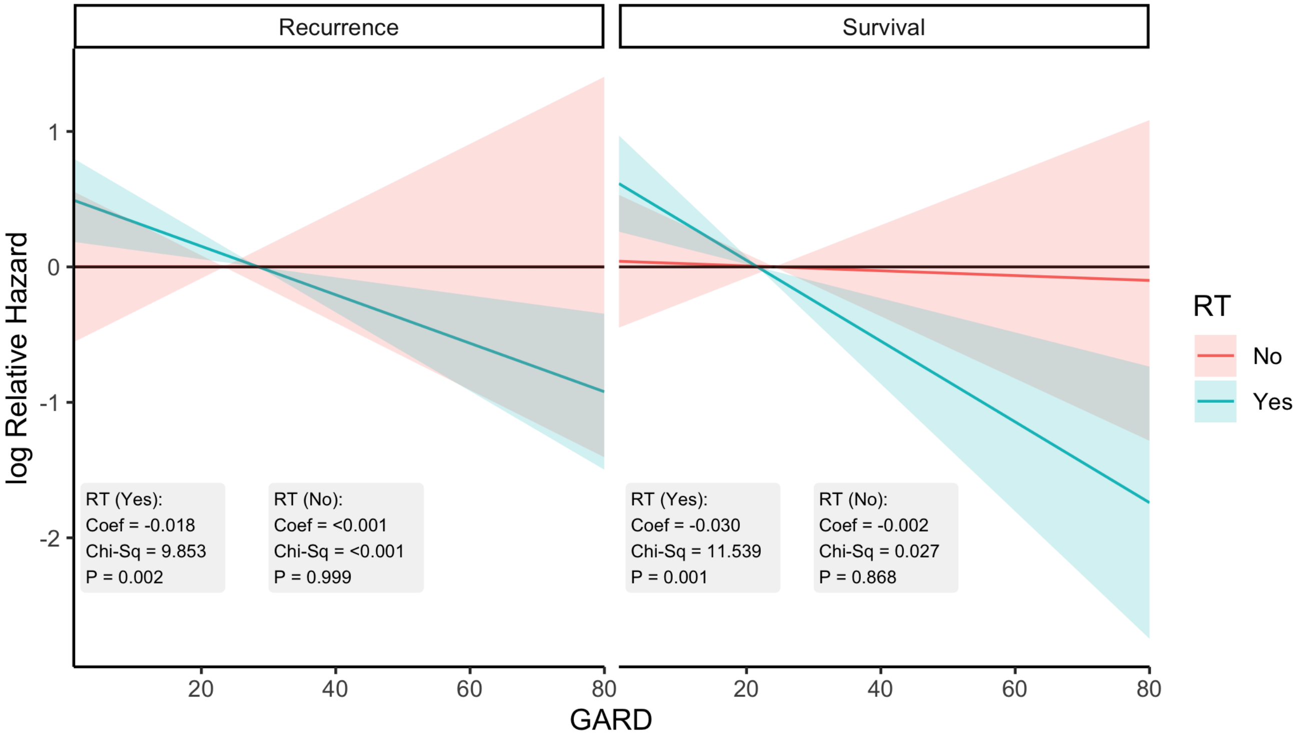


**Supplemental Figure 6. Stratified Cox regression analysis of HR for first recurrence (left) and overall survival (right) as function of GARD.** First recurrence for radiation treated patients, **left**, is significantly associated with GARD (coefficient $= -0.018$, $\chi$-squared $= 9.9$, $p = 0.002$), and is not for patients not treated with radiation (sham-GARD, coefficient $< 0.001$, $\chi$-squared $< 0.001$, $p = 0.999$). Overall survival for radiation treated patients, **right**, is significantly associated with GARD (coefficient $= -0.030$, $\chi$-squared 11.5, $p = 0.001$), and is not for patients not treated with radiation (sham-GARD, coefficient $= -0.002$, $\chi$-squared $< 0.01$, $p = 0.868$).

***Model residuals***

Model residuals for each of the 4 conditions (+/- RT, OS/Recurrence) are presented in **Supplemental Figure 7**.

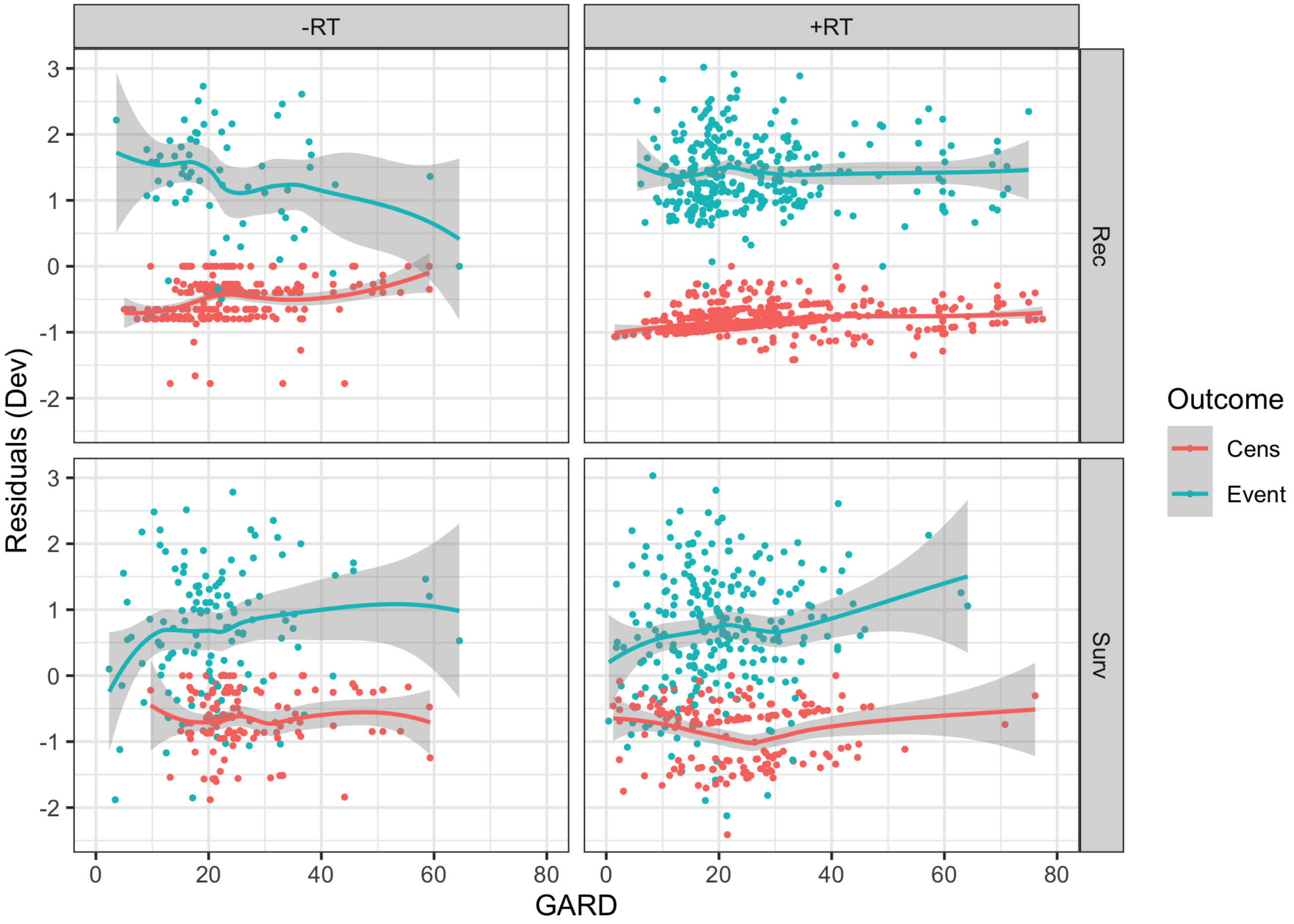


**Supplemental Figure 7. Deviance residuals for each of the stratified Cox regression models on GARD, for the no-RT (left) and with-RT (right) groups, using first recurrence (top) and overall survival (bottom).**
